# Health Economic Burden of COVID-19 in Saudi Arabia

**DOI:** 10.1101/2022.04.08.22273439

**Authors:** Khalidah A. Alenzi, Hamdan S. Al-malky, Ali F. Altebainawi, Hamidah Q. Abushomi, Fahad O. Alatawi, Moosa H. Atwadi, Moosa A. Khobrani, Dlal A. Almazrou, Nariman Alrubeh, Zainab A. Alsoliabi, Abdulaziz M. Kardam, Shakr A. Alghamdi, Abdulrahman Alasiri, Thamir M. Alshammari

**Affiliations:** Regional Drug Information Center, Ministry of Health, Tabuk, Saudi Arabia; Regional Drug Information Center, Ministry of Health, Jeddah, Saudi Arabia; Pharmaceutical Care Services, King Salman Specialist Hospital, Hail Health Cluster, Ministry of Health, Hail, Saudi Arabia; Medication Safety Research Chair, King Saud University, Riyadh, Saudi Arabia; Dammam medical complex, Eastern region cluster, Dammam, Saudi Arabia; King Fahad Specialist hospital, Ministry of Health, Tabuk, Saudi Arabia; Althager General Hospital, Ministry of Health, Jeddah, Saudi Arabia; Prince Mohammed bin Nasser Hospital, Ministry of Health, Jizan, Saudi Arabia; King Saud Medical City, Ministry of Health, Riyadh, Saudi Arabia; Qatif Central Hospital, Ministry of Health, Eastern Region, Saudi Arabia; King Faisal Medical City, Ministry of Health, Abha, Saudi Arabia; Khamis Mushait General Hospital, Ministry of Health, Southern Region, Saudi Arabia; Mohayl General Hospital, Ministry of Health, Asir, Saudi Arabia; College of Applied Medical Sciences, King Saud University, Riyadh, Saudi Arabia

## Abstract

**Background:** The coronavirus disease 2019 (COVID-19) pandemic has placed a massive economic burden on health care systems worldwide. Saudi Arabia is one of the numerous countries that have been economically affected by this pandemic. The objective of this study was to provide real-world data on the health economic burden of COVID-19 on the Saudi health sector and assess the direct medical costs associated with the management of COVID-19.

**Methods:** A retrospective cohort study was conducted based on data collected from patients hospitalized with COVID-19 across ten institutions in eight different regions in Saudi Arabia. The study calculated the estimated costs of all cases during the study period by using direct medical costs. These costs included costs directly related to medical services, such as the health care treatment, hospital stays, laboratory investigations, treatment, outcome, and other related care.

**Results:** A total of 5,286 adult patients admitted with COVID-19 during the study period were included in the study. The average age of the patients was 54 years, and the majority were male. Among the COVID-19 patients hospitalized in a general ward, the median hospital length of stay was 5.5 days (mean: 9.18 days), while the ICU stay was 4.26 days (mean: 7.94 days). The total medical costs for general ward and ICU patients were 14,585,640 SAR and 90,776,250 SAR, respectively. The total laboratory investigations ranked as the highest-cost services (22,086,296 SAR), followed by treatment (14,574,233.1 SAR). Overall, the total cost of all medical services for patients hospitalized with COVID-19 was 193,394,103.1 SAR.

**Conclusion:** This national study found that COVID-19 was not only a serious concern for patients but also a serious economic burden on the health care system in Saudi Arabia.

**Key points:**

1. The nursing costs and length of stay were lower in the ICU than in the general ward.
2. The costs of hospitalization in general medical wards were less than those of admission to the ICU.
3. These cost data will be valuable for future researchers evaluating the COVID-19 pandemic’s increasing health care economic burden in Saudi Arabia and the implementation of cost-effective models to assess the possible implications of COVID-19 prevention and treatment initiatives.

## Introduction

Coronavirus disease 2019 (COVID-19) is a respiratory infection caused by severe acute respiratory syndrome coronavirus 2 (SARS-CoV-2), which originated in Wuhan, Hubei Province, China, in December 2019^1,2^. COVID-19 was confirmed as a pandemic virus-related infection by the World Health Organization (WHO) in March 2020^3^.

Following the WHO pronouncement, countries worldwide, including the Kingdom of Saudi Arabia (KSA), began focusing on pandemic response plans to combat the spread of the virus. Since the first Saudi Arabian case of COVID-19 was confirmed on March 2, 2020, the KSA has implemented many measures to combat the spread of the disease^4^.

KSA is home to 35 million people, the majority of whom live in cities (84%), and the country experienced severe pressure on urban hospitals during the peak of the epidemic^5^. The Ministry of Health (MoH) of Saudi Arabia oversees most health care operations and services in the kingdom. In Saudi Arabia, the Ministry of Health has played a key role in offering health care services, including precautionary, therapeutic, and rehabilitation efforts^6, 7^. The government has endeavored to strengthen the health system and accelerate a health care transition by developing Public Private Participation (PPP) health care models, with the goal of increasing private sector engagement in overall health care spending to 35% by the year 2020^8^. In 2018, the MoH provided 58.3% of all hospitals and 59.1% of all beds in the nation^9^.

Saudi Arabia has announced a 32 million USD intervention to help economic sectors impacted by COVID-19. The debt ceiling was increased from 30% to 50% of GDP, while fiscal debt was foreseen to increase from 6.4% to 9% of GDP ^10^.

A major concern regarding the COVID-19 pandemic is the high-cost burden on health care systems. A study by Khan et al. calculated the direct medical costs associated with the treatment of COVID-19 patients in Saudi Arabia^11^. Based on the degree of care and length of stay, the total direct medical costs per patient were estimated. The total direct medical expenditure per patient hospitalized in the general medical ward (GMW) for moderate-to-severe symptoms was SAR 42,704.49 (US$ 1 = SAR 3.75). In contrast, intensive care unit (ICU) patients generated an approximately threefold cost increase (e.g., SAR 79,418.30). Surprisingly, the overall cost of care for patients on mechanical ventilators (MVs) was slightly lower than that for patients admitted to the GMW but not on MVs. This difference was mostly due to patients on MVs having a much shorter period of survival and higher mortality rate, which resulted in a shorter length of stay (LOS) and, subsequently, a lower overall cost per patient^12^.

According to projections, the total direct medical cost in the United States has ranged from US$ 163.4 billion to US$ 654.0 billion over the course of the pandemic^13^. In Sweden, the total direct medical cost has been projected to reach US$ 2 billion^14^. The calculated mean direct medical cost per patient was observed to be SAR 48,436.18 (US$ 12,916.31), which was not dramatically different from the cost published in Saudi Arabia for the management of MERS-CoV patients (US$ 12,947.03)^15^.

There is a scarcity of data on the direct medical costs of COVID-19 worldwide. Therefore, this study was conducted to present evidence-based statistics on COVID-19’s health economic burden on the Saudi health care system.

The study aims to provide real-world data on the health economic burden of COVID-19 on the Saudi health sector and assess the direct medical costs associated with the management of COVID-19.

## Methods

### Study design and data source

A retrospective cohort study was conducted based on data collected on patients hospitalized with COVID-19 at ten institutions in nine different regions in Saudi Arabia. The study period started on March 1st, 2020, and ended on January 30th, 2021. The patients included in this study were individuals who were hospitalized and followed until discharge or in-hospital death or those whose final follow-up event occurred at the latest on January 30th, 2021.

The study calculated the economic costs of all cases from the beginning of March to the end of January 2020 using the micro-cost method. The costs included in the study were the direct medical costs, i.e., the costs directly related to medical services, such as health care services, hospital stays, laboratory investigations, treatment, outcomes, and other related care.

The following variables were collected from patient medical records and anonymously entered into a web-based electronic form. When possible, the patient data included demographics characteristics (e.g., age, gender, marital status, and geographic region). Medical information (e.g., comorbidities, length of hospital stay, length of intensive care unit or isolation room or general ward stay) was also collected. Furthermore, the direct medical care costs were collected, including laboratory and diagnostic test costs (e.g., complete blood count (CBC), liver and cardiac enzymes, renal functions, biochemistry, swabs, cultures, chest X-rays, and computerised tomography (CT) scans), treatment costs including medications (e.g., antivirals, antimalaria, biologics, antibiotics, immunoglobulin, anticoagulants, and plasma), supportive therapy (e.g., mechanical ventilation and pneumatic compression devices), hospital stay (including room fees; intravenous sets and fluids in isolation rooms, intensive care units, and general wards; and cost of care provided by physicians and nurses).

All medications were obtained from the National Unified Procurement Company pricing list for medications that are available according to the Ministry of Health (antivirals, antimalarial drugs, biologics, antibiotics, immunoglobulin, anticoagulants). In addition, other medical costs, such as hospitalization fees (including for general wards and intensive care units (ICUs)) and fees for physician consultations and nurse care, laboratory tests, diagnostics tests, and all supportive therapies (mechanical ventilation, intravenous fluids, and pneumatic compression devices), were obtained from the Ministry of Health.

### Excluded costs

Personal protective equipment (e.g., N95 masks, gowns, protective eyewear), oxygen, plasma therapy, over-the-counter medicines, and the burden-increasing cost of comorbid diseases.

### Statistical analyses

The data are presented using descriptive statistics (mean, frequencies, and percentages). All analyses were conducted using statistical analysis software (SAS®; version 9.4, SAS Institute Inc., Cary, NC, USA).

## Results

### Epidemiological findings

A total of 5,286 adult patients admitted with COVID-19 during the period March 2020 to January 2021 were included in the study. As shown in Figure 1, the Eastern region of Saudi Arabia had the highest rate of admitted COVID-19 patients (21.69%), followed by Asir (19.73%). The lowest rates were found for the Al-Qassim region and Medina region.

**Figure 1:**
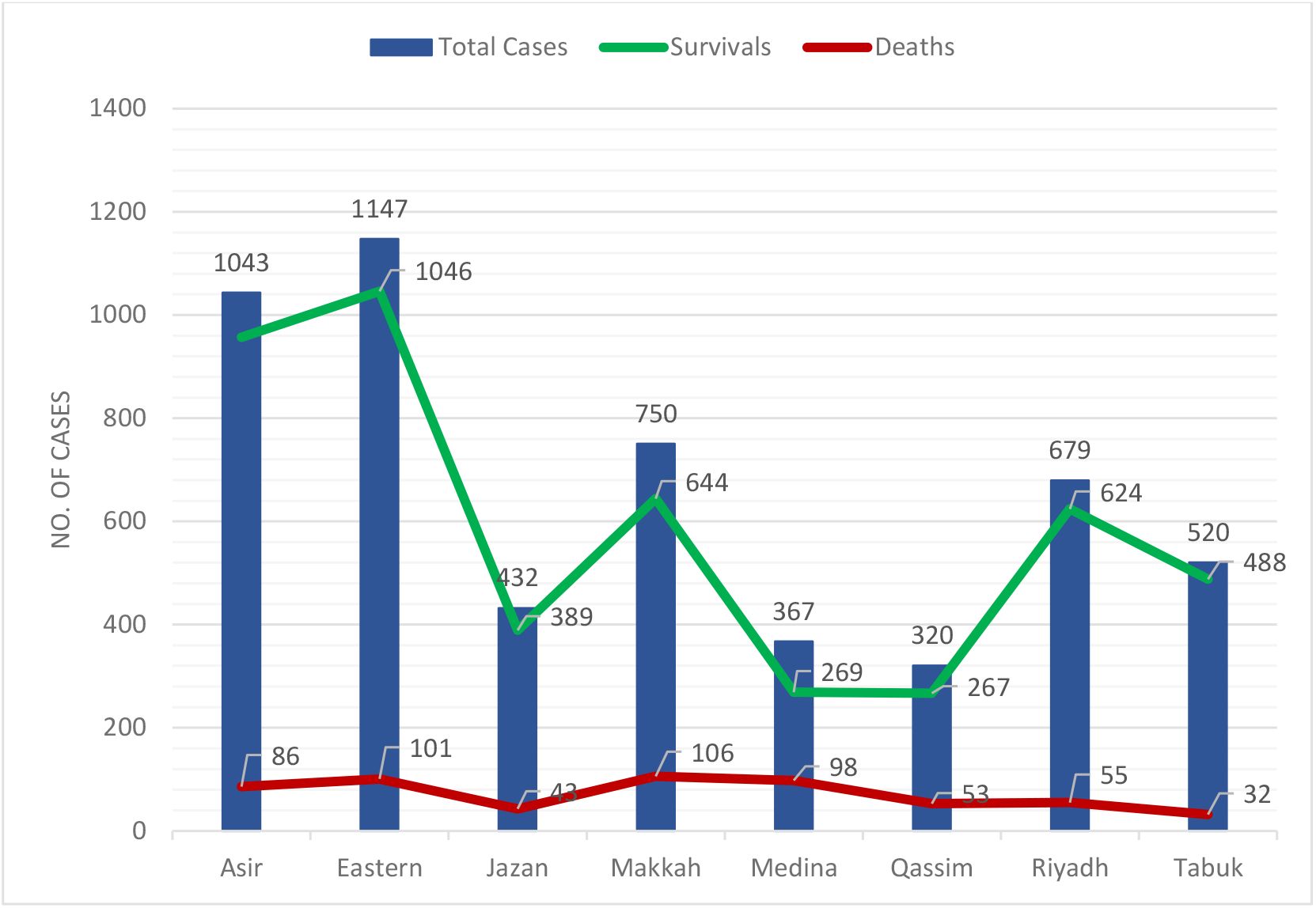
COVID-19 discharge and death rates among Saudi Regions, Mar 2020 - Jan 2021.

The patients’ baseline demographic and general characteristics are shown in Table 1. The age of the participants ranged from 18 years to more than 65 years, whereby males comprised the majority of the study population (79%). The largest proportion of patients consisted of those aged 45-65 years (approximately 44.32%), followed by patients over 65 years (25.50%).

**Table 1:** Baseline Characteristics of Patients Hospitalized With COVID-19.

| Variables | Relationship with death and discharge |  |  |  | Total |  |
| --- | --- | --- | --- | --- | --- | --- |
|  | Discharge (N=4,712) |  | Death (N=574) |  | N = 5286 | % |
|  | N | % | N | % |  |  |
| Groups of age (years) |  |  |  |  |  |  |
| 18 - 29 | 241 | 4.56 | 15 | 0.28 | 256 | 4.89 |
| 30 - 44 | 1190 | 22.51 | 102 | 1.93 | 1292 | 24.44 |
| 45 - 64 | 2091 | 39.56 | 252 | 4.77 | 2343 | 44.32 |
| ≥ 65 | 1146 | 21.68 | 202 | 3.82 | 1348 | 25.50 |
| Gender |  |  |  |  |  |  |
| Male | 3688 | 69.77 | 493 | 9.33 | 4181 | 79.1 |
| Female | 1028 | 19.45 | 81 | 1.53 | 1109 | 20.98 |
| Smoking | 1727 | 32.67 | 283 | 5.35 | 2010 | 38.02 |
| Supporting Therapy |  |  |  |  |  |  |
| Mechanical ventilation | 1554 | 29.4 | 218 | 4.12 | 1772 | 33.52 |
| Pneumatic compression devises | 230 | 4.35 | 2 | 0.04 | 232 | 4.39 |
| Comorbidities | 2940 | 55.62 | 496 | 9.4 | 3436 | 65 |
| Hypertension | 1527 | 28.89 | 198 | 3.74 | 1725 | 32.6 |
| Diabetes mellitus | 1388 | 26.26 | 253 | 4.79 | 1641 | 31.04 |
| Heart disease | 328 | 6.21 | 116 | 2.19 | 444 | 8.4 |
| Asthma | 233 | 4.41 | 22 | 0.41 | 255 | 4.82 |
| Cancer | 33 | 0.62 | 46 | 0.87 | 79 | 1.5 |
| Kidney disease | 112 | 2.12 | 23 | 0.44 | 135 | 2.56 |
| Obese | 183 | 3.46 | 23 | 0.44 | 206 | 3.89 |
| Anemia | 75 | 1.42 | 2 | 0.04 | 77 | 1.46 |
| Benign prostatic hyperplasia | 54 | 1.02 | 12 | 0.23 | 66 | 1.25 |
| Ischemic stroke | 52 | 0.98 | 30 | 0.57 | 82 | 1.55 |
| Epilepsy | 47 | 0.89 | 17 | 0.32 | 64 | 1.21 |
| Autoimmune disease | 31 | 0.59 | 3 | 0.06 | 34 | 0.63 |

Approximately 89.14% of the patients were discharged from the hospital, whereas 10.85% died. The mortality rate was higher among males (85.88%) than among females (14.12%) (Table 1).

Among the COVID-19 patients hospitalized in general wards, the median hospital length of stay (LOS) was 5.5 days (mean: 9.18 days), while the median ICU stay was 4.26 days (mean: 7.94 days) (Figure 2).

**Figure 2:**
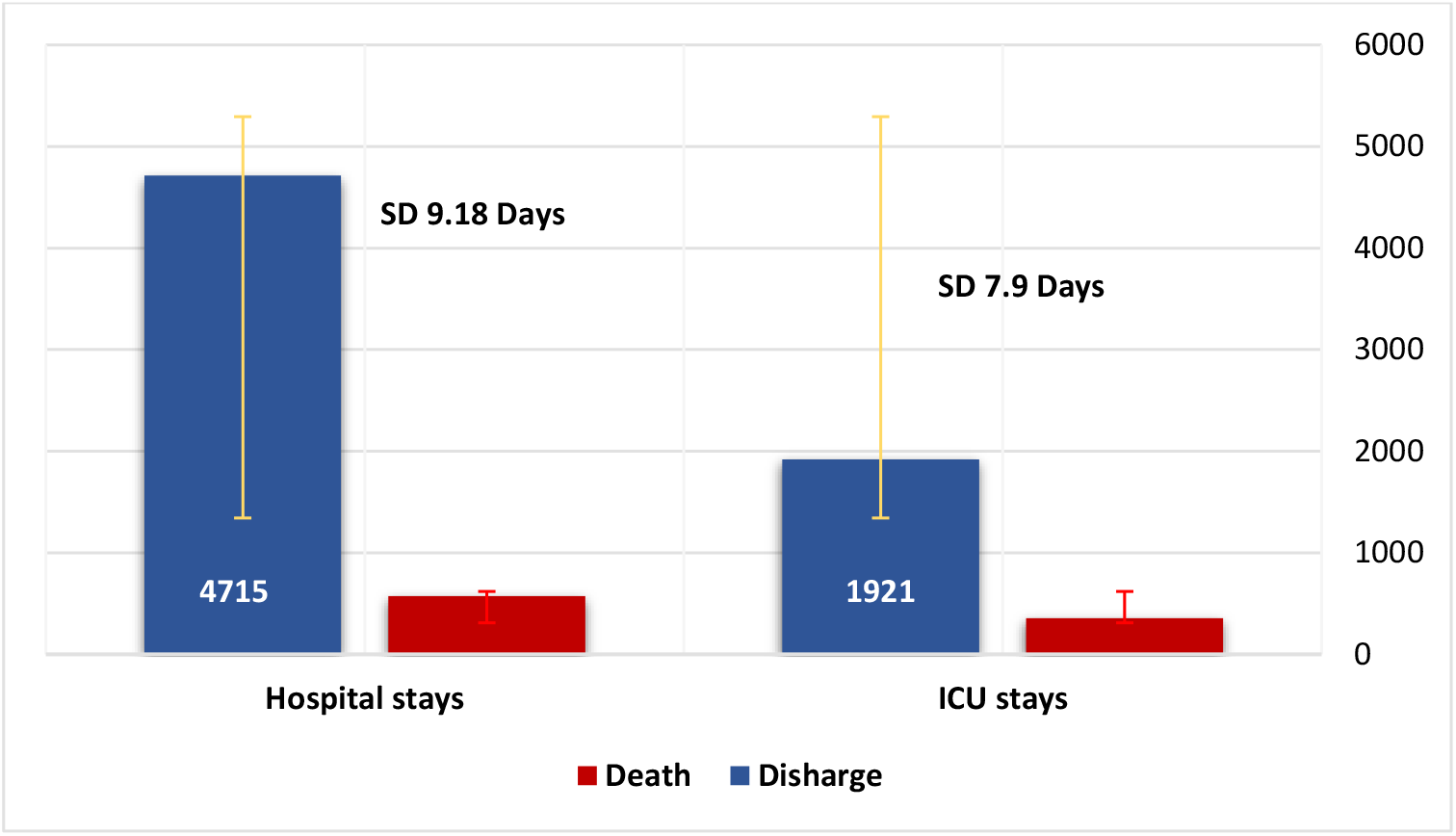
Hospital and ICU stay among discharged and non-surviving patients.

A relatively high number of patients (41.90%) were admitted to the intensive care unit .Of these, 33.52% used mechanical ventilation, and 78.40% of these patients had comorbidities.

The most common drug provided to patients was dexamethasone (44.82%), followed by favipiravir (39.29%) and hydroxychloroquine (26.63%). All patients were given more than one prophylactic antibiotic, and ceftriaxone was the most prescribed antibiotic (92.87%), followed by azithromycin (64.55%).

The total cost of general ward, isolation room and ICU stays was 14,585,640 SAR, 17,534,297.08 SAR, and 90,776,250 SAR, respectively, including room fees and intravenous sets and fluids (Table 2). Additionally, nursing care costs amounted to 12,134,320 SAR (7,292,820 SAR for general-ward care and 4,841,500 SAR for ICU care). The total physician consultation cost (12,134,680 SAR) was divided into ICU (4,841,860 SAR) and general-ward as well as isolation room (7,292,820 SAR) costs.

**Table 2:** Cost of hospital services for COVID-19 patients:

|  | Total | Median [IQR] | Mean [SD] |
| --- | --- | --- | --- |
| Hospital stays | 122,896,187.1 | 3,050 | 27,019.23 |
| ICU stays | 90,776,250 | 41,250 | 29,804.08 |
| General ward stays | 14,585,640 | 2,700 | 3,674.73 |
| Isolation room stays | 17,534,297.08 | 650 | 5,881.62 |
| Total physician consultations | 12,134,680 | 1,850 | 1,985 |
| Total physician consultations -ICU | 4,841,860 | 2,700 | 1,589.55 |
| Total physician consultations – Ward & isolation | 7,292,820 | 1,350 | 1,837.37 |
| Total nursing care | 12,134,320 | 1,775 | 1,732.94 |
| ICU -Nursing care | 4,841,500 | 2,200 | 1,593 |
| Ward & isolation -Nursing care | 7,292,820 | 1,350 | 1,837.37 |
| Mechanical ventilation | 7,305,200 | 1,356.33 | 2,638.5 |
| Cuff of pneumatic compression device | 24,415 | 95 | 20.25 |
| Laboratory investigations | 22,086,296 | 335 | 356.6 |
| CBC | 557,376 | 192 | 67.91 |
| Liver enzymes | 1,620,800 | 300.93 | 184.8 |
| Cardiac enzymes | 4,167,360 | 240 | 1048.95 |
| Renal functions | 2,643,680 | 160 | 433.18 |
| Biochemistry | 1,629,120 | 480 | 240.88 |
| D-dimer | 1,651,360 | 560 | 465.79 |
| Swabs | 1,826,000 | 339.02 | 58.17 |
| Cultures | 984,232 | 476 | 286.36 |
| Antibiotic sensitivity test | 1,447,400 | 268.73 | 421.12 |
| Radiology services | 1,988,300 | 650 | 457 |
| Chest X-rays | 733,200 | 136.13 | 66.27 |
| CT scans | 1,255,100 | 1,100 | 504.63 |
| PCR | 5,810,000 | 1,050 | 185.08 |
| Medications | 14,574,233.03 | 2,705.45 | 1590 |
| Lopinavir/ritonavir | 68,293.12 | 47.25 | 32.26 |
| Favipiravir | 755,664 | 168 | 193.99 |
| Interferon beta-1 | 6,271,544.7 | 2,835.75 | 2988.16 |
| Hydroxychloroquine | 15,500 | 7 | 5.2 |
| Tocilizumab | 2,661,464.24 | 1,974.38 | 1,306.67 |
| Anticoagulants | 672,056.52 | 3,259.96 | 67.78 |
| Antibiotics | 2,132,708.87 | 8,977.28 | 185.37 |
| Immunoglobulin | 169,155 | 1,181.25 | 272.89 |
| Antifungals | 1,827,846.58 | 280 | 647.69 |
| Total cost | 193,394,103.1 | 16,655.13 | 44,315.77 |

CBC: Complete blood count , ICU: Intensive care unit , PCR :polymerase chain reaction.

Among the total hospital service costs, total laboratory investigations ranked the highest (22,086,296 SAR), followed by medications (14,574,233.1 SAR) (Table 2, Figure 3).

**Figure 3:**
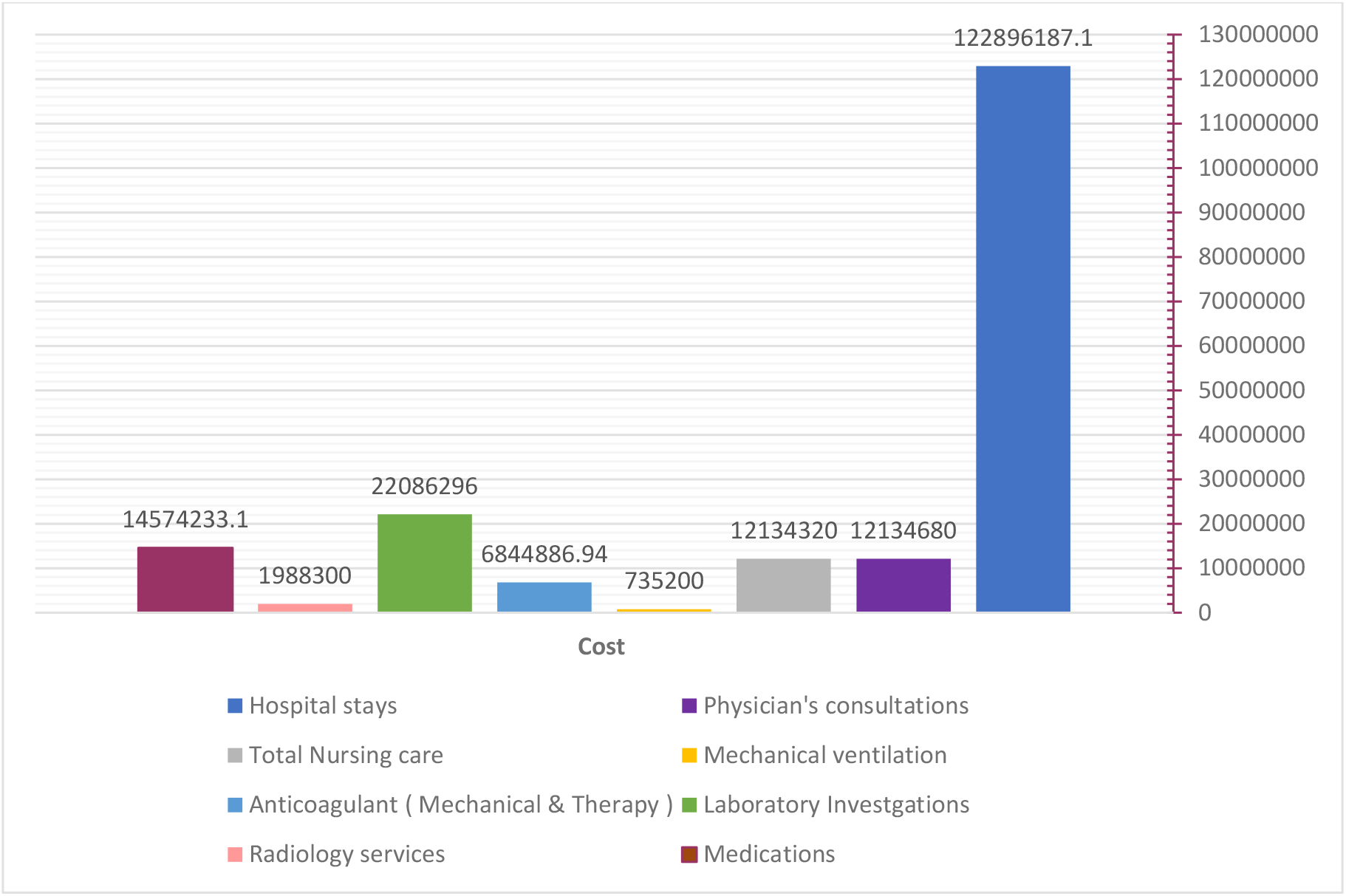
Distribution of Total Hospital Costs.

Additionally, the highest costs among the laboratory investigations were PCR testing (5,810,000 SAR), followed by cardiac enzymes investigations (4,167,360 SAR), while the majority of radiology service costs was for CT scans (1,255,100 SAR) (Table 2).

In addition, the total general-ward and isolation ward cost was 72,299,431.52 SR, and most of this cost was due to the stay cost (32,119,937.08 SAR), followed by laboratory investigations (14,289,816 SAR), medications (9716155.4 SAR), and radiology services (1,325,533.3 SAR) (Figure 4).

**Figure 4:**
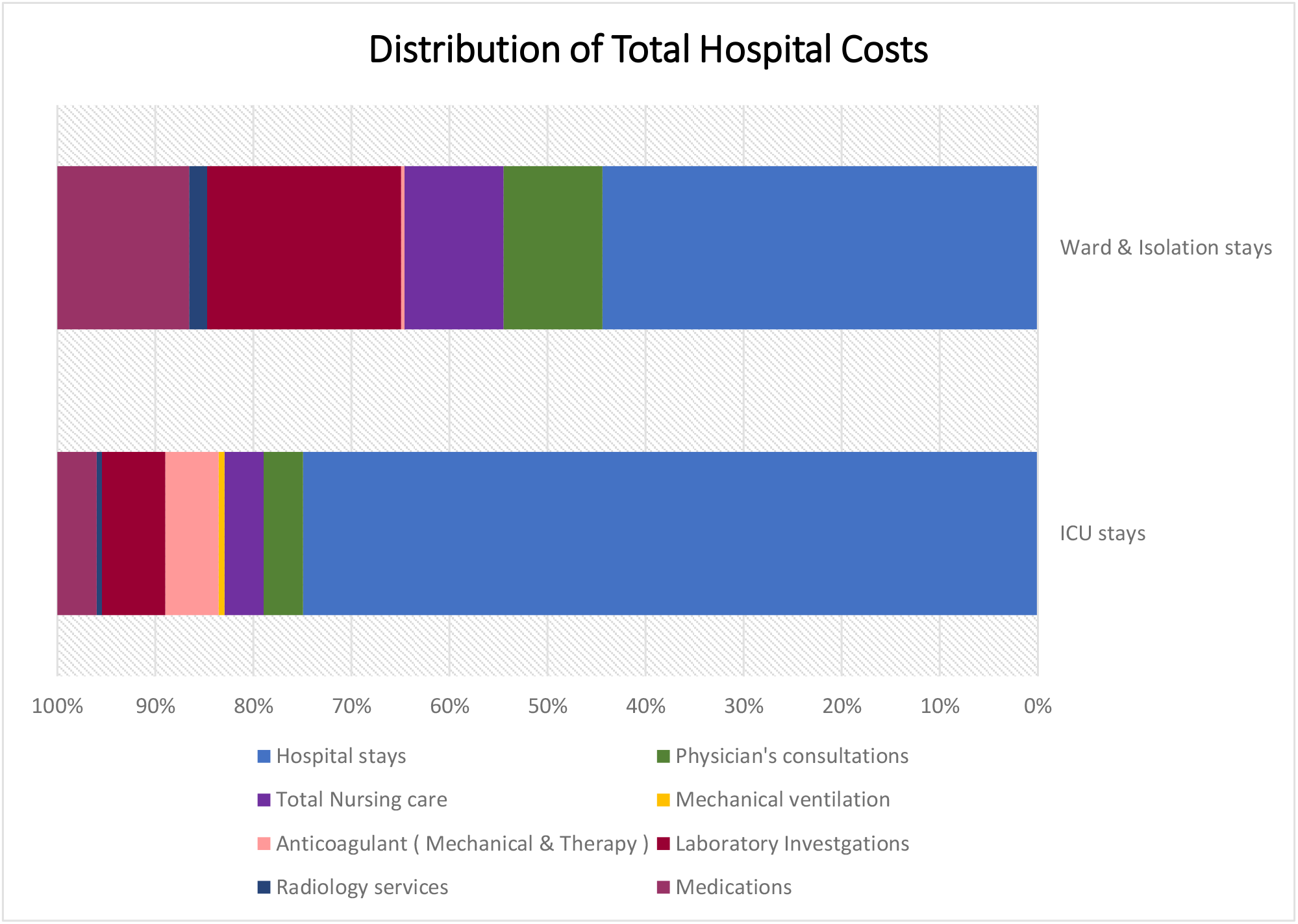
Distribution of Total Hospital Costs according to locations.

Among the total ICU service costs, total ICU laboratory investigations ranked the highest (7,796,480 SAR), followed by anticoagulant prophylaxis, including mechanical and anticoagulation drugs (6,586,770.2), and ICU medications (4,858,077.7 SAR) (Figure 4).

Interferon beta-1 was the highest-cost medication among the treatment options (6,271,544.7 SAR), followed by tocilizumab (2,661,464.24 SAR) and then antibiotics (2,132,708.87 SAR). The cost of each service, such as laboratory investigations and treatment per medication, is presented in Table 2 in detail.

In contrast, the total cost of patients who died due to COVID-19 was 64,464,701.03 SAR (mean: 15,257.962 SAR), while that of patients who recovered was 128,929,402.1 SAR (mean: 25,279 SAR) (Table 3).

**Table 3:** Distribution of Cost Components by Patient Outcomes.

|  | Death | Discharge |
| --- | --- | --- |
| Total Cost | 64,464,701.03 | 128,929,402.1 |
| Mean [SD] | 15,257.962 | 25,279 |

Overall, the total cost of all medical services for patients hospitalized with COVID-19 was 193,394,103.1 SAR (mean: 44,315.77 SAR) (Table 2).

## Discussion

Worldwide, health care systems have faced substantial difficulty in terms of resource consumption and expense management as a result of the COVID-19 outbreak. Governments and hospitals have been compelled to make challenging budget allocation decisions. For decision-making, empirical evidence is critical. To make accurate judgments, decision-makers could benefit from economic studies on COVID-19 medical treatment. This study contributes to a better understanding of medical care procedures and resource use. To the best of our knowledge, there have been only a few economic studies on COVID-19. However, we believe that our study provides more detailed information than other studies, as we calculated the cost for each item and service.

While differences in research methods, demographics, health-care costs, and other factors make it difficult to compare studies, particularly studies from different countries, several researchers have investigated the burden of health services on the health care system. All studies have found that the burden on the health care system in the form of resource use and expenses has been substantial.^16, 17-19^

This study determined that the total cost of 5,286 patients treated in different units for COVID-19 was 193,394,103.1 SAR (51,508,726.60 USD). Based on this outcome, the average cost per patient was determined to be ≈ 44,315.77 SAR (10,182.5 USD). The cost per patient has been reported to be 3,045 USD in the United States, 6,827 USD in China, 12,637.42 USD in Latin America, 2,192 USD in children aged 0-19 in Korea, 4,633.43 in India and 4,847 pounds sterling in the United Kingdom.^13, 16, 17, 20, 21^ These differences may be attributable to differing treatment regimens, preferences among health professionals, health care resource consumption rates, and medical equipment pricing levels from country to country.

In this study, it was observed that nursing costs and length of stay were lower in the ICU than in the general ward. This finding might be due to ICU patients having a much shorter period of survival and a higher risk of mortality, which frequently resulted in a shorter LOS and eventually death. However, because of the increased cost of health care resource utilization, the overall cost per patient in the ICU was substantially higher than that in the general ward. This observation is consistent with the findings of Rae et al.^12^

Moreover, in hospitalized patients in Turkey, the mean LOS was 8.0 days for patients hospitalized in general wards versus 14.8 days for patients hospitalized in ICUs. This finding is consistent with our study with respect to general-wards stays and significantly less than twice the LOS in the ICU^22^.

Moreover, our results found that the total ICU cost was 121,094,671.6 SAR (average cost ≈ 54,498.5 SAR), whereas the total general ward cost was 72,299,431.52 SAR (average cost ≈ 22,981.4 SAR), findings consistent with the results of a study by Khan AA et al. These findings reveal that the expense of medical care increases as the severity of the patient’s condition increases. However, the costs of hospitalization in general medical wards were less than those of admission to the ICU (42,704.49 SAR and 79,418.30 SAR, respectively)^11^. Notably, the Khan AA et al. study did not include detailed cost calculations and the total cost of all study populations. Additionally, they investigated fewer study populations. Moreover, ICU hospitalizations had the highest average daily cost, according to a study that only examined the direct medical costs of COVID-19 in South Africa.^23^ The increase in the cost of critical care may be due to most cases being severe with poor progression and (78.4%) involving comorbidities; most such cases required mechanical ventilation (78.4%) or expensive medications, such as tocilizumab, interferon beta-1 (according to the recommendations of the COVID-19 protocol from the MOH), or systemic antifungal medications.

Our study found that for most of the full hospital service costs, the hospital stay included room fees as well as cost of intravenous sets and fluids in isolation rooms, intensive care units, and general wards (63.55%), followed by laboratory investigations. The latter represented the highest cost (11.42%), while PCR tests were the most expensive among the laboratory tests (26.31%). A study conducted in Greece similarly founding the highest costs for RT–PCR tests and hospitalization^24^.

A Turkish study on 163 patients admitted to the ICU found that the largest share of ICU costs was associated with procedural packages (72%), followed by medication costs (12.8%). In our study, we found that the highest costs for patient care were associated with ICU stays, including procedural packages (75%) and laboratory investigations (6.44%), followed by anticoagulant prophylaxis, including mechanical and medications (5.44), and then treatment (4%)^22^.

The Ministry of Health in Saudi Arabia has released protocols to manage incidences despite the significant increases in morbidity and mortality from COVID-19 while considering drug treatment strategies for inpatients in addition to supporting medical resources and the limited use of medications, especially antibiotics. Our study found that antibiotics were the most commonly used medications in managing infections associated with COVID-19. The prescription of antibiotics peaked during the chaotic early days of the pandemic, when physicians did not know much about the virus and prescribed broad-spectrum antibiotics, which are very costly. Additionally, a number of medications were initially thought to be beneficial against COVID-19, such as favipiravir and hydroxychloroquine. However, studies have proven that these medications have no effect against COVID-19^25^. These developments represent additional reasons for the high cost of treating COVID-19 patients. In contrast, several expensive medications (e.g., remdesivir) were not utilized in high quantities during the study period, which might have lowered the treatment cost.

Subsequently, the ministry has issued an updated therapeutic protocol based on global evidence regarding the effectiveness and safety of the medications noted above. As all countries of the world have adopted similar guidelines, the consumption of these medicines has increased. However, the COVID-19 epidemic has caused significant supply chain disruptions and pricing increases for local raw materials, which have disproportionately impacted smaller markets and increased the costs of medications in markets with increased consumption^26^. In Denmark, price increases for versions of ten medications averaged 71.6 percent. The increases averaged 43% and 37% in the Netherlands and Sweden, respectively. Price increases up to 49% were observed in the United Kingdom, primarily for azithromycin. This burden has also affected the Saudi market ^26^.

The expenses of medical treatment for COVID-19 are not only a concern for hospitals. The expense of COVID-19 is placing a strain on a number of government agencies. Regardless whether care is provided in private hospitals, the Ministry of Health covers all costs^6^. Medical expenditures are increasing in line with the severity of cases, placing greater strain on health care systems. Therefore, this research may be of assistance to all governing bodies in planning activities and making choices in connection with the pandemic because it continues to affect all countries and because the study is specific to Saudi Arabia. Additionally, it could help in planning for any future pandemic or other outbreak of communicable and noncommunicable disease.

This study has several strengths and limitations. Two limitations are as follows. 1) Because the data were collected from patient medical records, certain information was unavailable, such as the frequencies of laboratory and radiology services. 2) We could not determine the costs for personal protective equipment (e.g., N95 masks, gowns, protective eyewear), oxygen, plasma therapy, and over-the-counter medicines, or for the burden of comorbid disease.

However, the study has the advantage of being the first – to the best of our knowledge – to include a high number of patients and detailed cost information per item and service. Additionally, this study’s findings represent unique insight into the Ministry of Health Hospital’s economic burden while providing care for COVID-19-infected individuals in Saudi Arabia. These cost data will be valuable for future researchers evaluating the COVID-19 pandemic’s increasing health care economic burden in Saudi Arabia and the implementation of cost-effective models to assess the possible implications of COVID-19 prevention and treatment initiatives.

## Conclusion

This national study found that COVID-19 was not only a serious concern for patients but also a serious economic burden on the health care system in Saudi Arabia. This economic burden has affected all types of direct cost within the nation’s health institutions. The results of this study should be used to better allocate costs in future planning for pandemics or outbreaks of other diseases.

## Data Availability

All data produced in the present work are contained in the manuscript.

## References

1. Wu F, Zhao S, Yu B, Chen Y-M, Wang W, Song Z-G, et al. A new coronavirus associated with human respiratory disease in China. Nature. 2020;579:265–9.

2. Alamri F, Alsofayan Y, AlRuthia Y, Alahmari A, Almuzaini Y, Abo Gazalah F, et al. Predictors of Hospitalization Among Older Adults with COVID-19 in Saudi Arabia: A Cross-Sectional Study of a Nationally Representative Sample. Risk Manag Healthc Policy. 2021;14:875–86.

3. Chatterjee R, Ghosh M, Sahoo S, Padhi S, Misra N, Raina V, et al. Next-Generation Bioinformatics Approaches and Resources for Coronavirus Vaccine Discovery and Development-A Perspective Review. Vaccines. 2021;9.

4. Al-Hanawi MK, Angawi K, Alshareef N, Qattan AMN, Helmy HZ, Abudawood Y, et al. Knowledge, Attitude and Practice Toward COVID-19 Among the Public in the Kingdom of Saudi Arabia: A Cross-Sectional Study. Front public Heal. 2020;8:217.

5. Nicola M, Alsafi Z, Sohrabi C, Kerwan A, Al-Jabir A, Iosifidis C, et al. The socio-economic implications of the coronavirus pandemic (COVID-19): A review. Int J Surg. 2020;78:185–93.

6. Alshammari, T. M., Altebainawi, A. F., & Alenzi, K. A. (2020). Importance of early precautionary actions in avoiding the spread of COVID-19: Saudi Arabia as an Example. Saudi Pharmaceutical Journal, 28(7), 898–902.

7. Anas A. Khan, Yousef M. Alsofayan, Ahmed A. Alahmari, Jalal M. Alowais, Abdullah R. Algwizani, Haleema A. Alserehi, Abdullah M. Assiri HAJ. COVID-19 in Saudi Arabia: the national health response. East Mediterr Heal J. 2021;27:1114–23.

8. Rahman R, Qattan A. Vision 2030 and Sustainable Development: State Capacity to Revitalize the Healthcare System in Saudi Arabia. Inquiry. 2021;58:46958020984682.

9. Alumran A, Almutawa H, Alzain Z, Althumairi A, Khalid N. Comparing public and private hospitals’ service quality. J Public Health (Bangkok) [Internet]. 2021;29:839–45. Available from: https://doi.org/10.1007/s10389-019-01188-9.

10. Gulf Today. Saudi Arabia announces $32b support to mitigate coronavirus impact on economy [Internet]. 2020. [cited 2021 Dec 28]. Available from: https://www.gulftoday.ae/business/2020/03/20/saudi-arabia-announces-%2432b-support-to-mitigate-coronavirus-impact-on-economy

11. Khan AA, AlRuthia Y, Balkhi B, Alghadeer SM, Temsah M-H, Althunayyan SM, et al. Erratum: Khan, A.A., et al. Survival and Estimation of Direct Medical Costs of Hospitalized COVID-19 Patients in the Kingdom of Saudi Arabia (Short Title: COVID-19 Survival and Cost in Saudi Arabia). Int. J. Environ. Res. Public Health 2020, 17, 7458. Int. J. Environ. Res. Public Health. 2020.

12. Rae M., Claxton G., Kurani N., McDermott D. CC. Potential Costs of Coronavirus Treatment for People with Employer Coverage [Internet]. 2020 [cited 2021 Dec 28]. Available from: https://www.kff.org/coronavirus-covid-19/issue-brief/potential-costs-of-coronavirus-treatment-for-people-with-employer-coverage/

13. Bartsch SM, Ferguson MC, McKinnell JA, O’Shea KJ, Wedlock PT, Siegmund SS, et al. The Potential Health Care Costs And Resource Use Associated With COVID-19 In The United States. Health Aff (Millwood). United States; 2020;39:927–35.

14. Sjödin H, Johansson AF, Brännström Å, Farooq Z, Kriit HK, Wilder-Smith A, et al. COVID-19 healthcare demand and mortality in Sweden in response to non-pharmaceutical mitigation and suppression scenarios. Int J Epidemiol. 2020;49:1443–53.

15. AlRuthia Y, Somily AM, Alkhamali AS, Bahari OH, AlJuhani RJ, Alsenaidy M, et al. Estimation Of Direct Medical Costs Of Middle East Respiratory Syndrome Coronavirus Infection: A Single-Center Retrospective Chart Review Study. Infect Drug Resist. 2019;12:3463–73.

16. Li X-Z, Jin F, Zhang J-G, Deng Y-F, Shu W, Qin J-M, et al. Treatment of coronavirus disease 2019 in Shandong, China: a cost and affordability analysis. Infect Dis poverty. 2020;9:78.

17. Vernaz N, Agoritsas T, Calmy A, Gayet-Ageron A, Gold G, Perrier A, et al. Early experimental COVID-19 therapies: associations with length of hospital stay, mortality and related costs. Swiss Med Wkly. Switzerland; 2020;150:w20446.

18. Gedik H. THE COST ANALYSIS OF INPATIENTS WITH COVID-19. Acta Medica Cordoba. 2020;36:3289

19. Lee JK, Kwak BO, Choi JH, Choi EH, Kim JH, Kim DH. Financial Burden of Hospitalization of Children with Coronavirus Disease 2019 under the National Health Insurance Service in Korea. J Korean Med Sci. 2020;35:e224.

20. R SR. Rs 3.5 lakh spent on each Covid-19 patient in Victoria Hospital: Karnataka minister [Internet]. 2020 [cited 2021 Dec 28]. Available from: https://timesofindia.indiatimes.com/city/bengaluru/rs-3-5-lakh-spent-on-each-covid-19-patient-in-victoria-hospital-karnataka-minister/articleshow/75639482.cms

21. Thom H, Walker J, Vickerman P, Hollingworth W. Exploratory comparison of Healthcare costs and benefits of the UK’s Covid-19 response with four European countries. Eur J Public Health. 2021;31:619–24.

22. Oksuz E, Malhan S, Gonen MS, Kutlubay Z, Keskindemirci Y, Tabak F. COVID-19 healthcare cost and length of hospital stay in Turkey: retrospective analysis from the first peak of the pandemic. Health Econ Rev. 2021;11:39.

23. Edoka I, Fraser H, Jamieson L, Meyer-Rath G, Mdewa W. Inpatient Care Costs of COVID-19 in South Africa’s Public Healthcare System. Int J Heal policy Manag. Iran; 2021;

24. Maltezou HC, Giannouchos T V, Pavli A, Tsonou P, Dedoukou X, Tseroni M, et al. Costs associated with COVID-19 in healthcare personnel in Greece: a cost-of-illness analysis. J Hosp Infect. 2021;114:126–33.

25. Bosaeed, M., Alharbi, A., Mahmoud, E., Alrehily, S., Bahlaq, M., Gaifer, Z., … & Alaskar, A. (2022). Efficacy of favipiravir in adults with mild COVID-19: a randomized, double-blind, multicenter, placebo-controlled trial clinical trial. Clinical Microbiology and Infection.

26. Ando G. Prices of essential COVID-19 medicines have increased 4% globally since February. [Internet]. 2020. [cited 2021 Dec 28]. Available from: https://www.pharmaceutical-technology.com/pricing-and-market-access/prices-essential-covid19-medicines-increased-4-percent-globally-html/

